# Seattle Flu Study - Swab and Send: Study Protocol for At-Home Surveillance Methods to Estimate the Burden of Respiratory Pathogens on a City-Wide Scale

**DOI:** 10.1101/2020.03.04.20031211

**Authors:** Ashley E. Kim, Elisabeth Brandstetter, Chelsey Graham, Jessica Heimonen, Audrey Osterbind, Denise J. McCulloch, Peter D. Han, Lea M. Starita, Deborah A. Nickerson, Margaret M. Van de Loo, Jennifer Mooney, Mark J. Rieder, Misja Ilcisin, Kairsten A. Fay, Jover Lee, Thomas R. Sibley, Trevor Bedford, Janet A. Englund, Michael Boeckh, Helen Y. Chu, on behalf of the Seattle Flu Study Investigators

**Author notes:** For complete investigator list, see below (on behalf of the Seattle Flu Study Investigators).

## Abstract

**Introduction:** While seasonal influenza and other respiratory pathogens cause significant morbidity and mortality each year, the community-based burden of these infections remains incompletely understood. Understanding the prevalence, epidemiology, and transmission dynamics of respiratory pathogen infections among community-dwelling individuals is essential during pandemic and epidemic settings and for developing pandemic-preparedness infrastructure.

**Methods and Analysis:** We present the protocol for a novel, city-wide home-based cross-sectional study in the Seattle Metropolitan area, utilizing rapid delivery systems for self-collection of a nasal swab and return to the laboratory for respiratory pathogen testing. All participation takes place electronically, including recruitment, consent, and data collection. Within 48 hours of participants self-reporting respiratory symptoms, a nasal swab kit is delivered to the household via a courier service. Demographic and illness characteristics are collected at the time of sample collection and recovery and behavioral information collected one week later. Specimens are tested in the laboratory for multiple respiratory pathogens, and results are available on a public website for participants.

**Ethics and Dissemination:** The study was approved by the University of Washington Institutional Review Board (Protocol #00006181). Results will be disseminated through peer-reviewed publications, talks at conferences, and on the *Study Website* (www.seattleflu.org).

**Article Summary:**

- The findings of this study will inform whether a home-based approach to city-wide respiratory surveillance is possible in epidemic settings
- A key strength of this study is that it is conducted across diverse neighborhoods spanning a major metropolitan area
- Clinical findings may not be generalizable, as they are reliant on self-report (vaccination status, symptoms, healthcare utilization, etc.)

## Introduction

Acute respiratory illnesses (ARIs) pose a significant burden on the healthcare system in the United States and represent an important cause of morbidity and mortality worldwide[1-4]. In the United States, influenza is estimated to cause 9-45 million illnesses 140,000-810,000 hospitalizations, and 12,000-67,000 deaths while respiratory syncytial virus (RSV) is estimated to cause 2 million outpatient visits each year in children under the age of 5[5,6]. Despite this burden, estimates about prevalence of the pathogens that result in ARIs rely on in-person healthcare visits or on aggregate counts from hospitalized individuals[6-10]. Additionally, these estimates generally capture illnesses when specific symptoms are present, such as fever, cough, or sore throat. Thus, they are likely a poor proxy for the prevalence of illness in community-dwelling individuals who may not necessarily seek care for their illness or have symptoms that fall outside of these criteria.

Active, community-level surveillance for respiratory pathogens is essential to monitor the seasonal activity of these pathogens, as it may inform public health prevention strategies or influence treatment decisions made at the community level. Previous studies focused on respiratory pathogen surveillance only studied specific subsets of the population, such as households with children, or required a significant, coordinated effort to capture the community, which makes the approach difficult to replicate[11-13].

Additionally, similar to traditional respiratory surveillance networks, these studies relied on in-person clinic visits which may contribute to potential spread of these pathogens. Nevertheless, community-wide surveillance studies provide an opportunity to better understand these pathogens among symptomatic individuals with wide-ranging illness severities and variable healthcare-seeking behaviors.

This manuscript describes the protocol for the Seattle Flu Study (SFS) Swab and Send sub-study, a novel, city-wide cross-sectional study of respiratory pathogens that yielded community-level surveillance through a home-based approach. The objectives of this study are to gather data from the community to more accurately estimate the prevalence of respiratory illness, to examine whether an exclusively online and home-based mechanism for data collection is feasible in an epidemic setting, and to determine whether these methods are scalable in the context of a pandemic.

### Aims and Hypothesis

The primary aim of this study is to assess the feasibility of a novel, city-wide home-based cross-sectional study in the Seattle Metropolitan area, utilizing rapid delivery systems for self-collection of a nasal swab from individuals experiencing a new ARI with return of specimens to the laboratory for respiratory pathogen testing.

We hypothesize that novel, home-based surveillance of influenza and other respiratory pathogens can be utilized to monitor the epidemiology and transmission dynamics of influenza and other respiratory pathogens at a city-wide scale in Seattle, Washington, USA.

## Methods and Analysis

### Study Design

This is a prospective study of individuals residing within the Seattle Metropolitan area with ARI symptoms. The study is nested within the Seattle Flu Study, a multi-armed influenza surveillance mechanism described, the protocol of which is described elsewhere[14].

### Study population inclusion/exclusion criteria

Individuals are eligible to participate in this study if they live within the Seattle Metropolitan area, experience a new or worsening cough and/or two ARI symptoms in the past seven days at time of enrollment (Table 1), are English-speaking, and have a valid email address and access to the internet at home. All individuals are consented to participate in the research study electronically. A parent or legally-authorized representative (LAR) will consent for individuals under 18 years, with assent for those between 7-18 years (Figure 1).

**Table 1.** List of symptoms that will be used in online questionnaires to screen individuals for eligibility. Selecting either acute cough *or* two or more concurrent *qualifying* symptoms will be considered an acute illness episode and will make an individual eligible for enrollment in the Swab and Send study.

|  |  |
| --- | --- |
| Feeling feverish or warm * | Runny/stuffy nose or sneezing * |
| Headache * | Feeling more tired than usual * |
| New or worsening cough ** | Muscle or body aches * |
| Chills or shivering <sup>x</sup> | Increased trouble with breathing * |
| Sweats <sup>x</sup> | Diarrhea <sup>+</sup> |
| Sore throat or itchy/scratchy throat * | Ear pain/ear discharge <sup>+</sup> |
| Nausea or vomiting * | Rash <sup>+</sup> |

\* a qualifying symptom for study eligibility for individuals of any age
\*\* a qualifying symptom that is sufficient on its own for study eligibility for individuals of any age
<sup>x</sup> not a qualifying symptom for study eligibility
<sup>+</sup> a qualifying symptom for study eligibility for individuals <18 years of age

**Figure 1.**
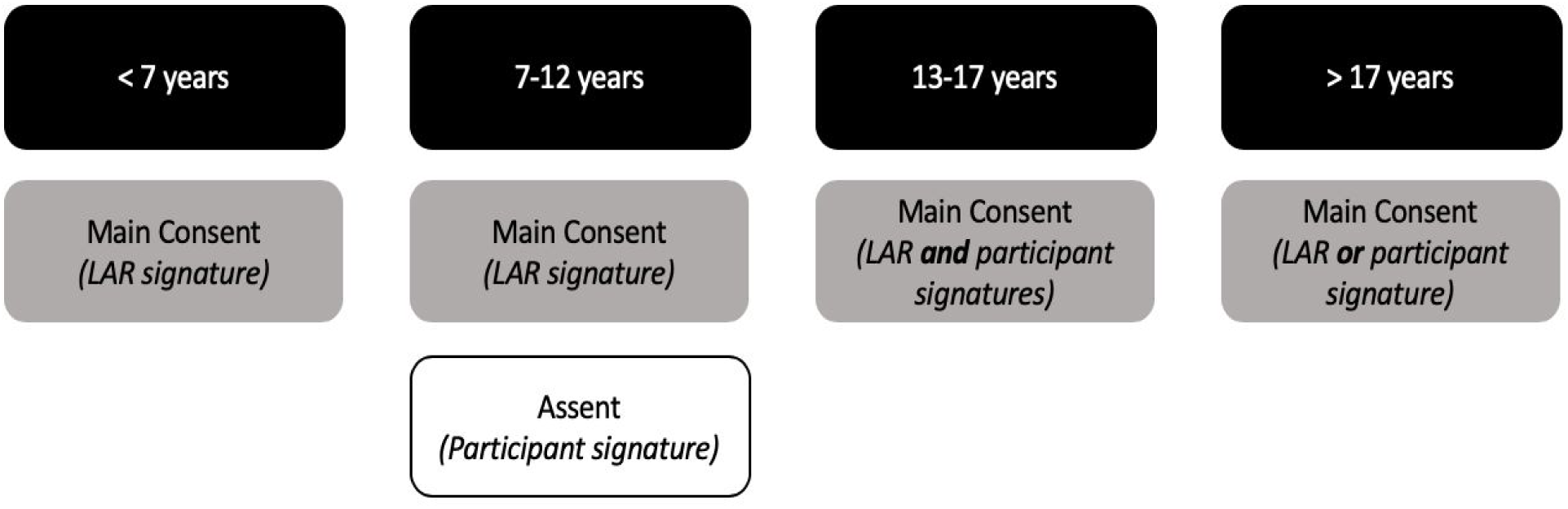
Documentation of written, informed consent by age of participant in the community cross-sectional, and prospective clinical and childcare cohorts of the Seattle Flu Study. If the participant is unable to provide informed consent due to cognitive impairment or because they have not attained the legal age for consent, a legally-authorized representative (LAR) may sign the consent form on their behalf. Participants enrolled by Seattle Children’s Hospital staff or participants enrolled into the prospective clinical cohort sign a HIPAA Agreement in addition to the main consent.

Individuals are excluded from participation if they are incarcerated, wards of the state, unable to comply with study procedures at the investigator’s discretion, or previously enrolled in Swab & Send in the past seven days.

### Recruitment, Screening, and Consent

Recruitment of potential study participants is conducted using a combination of in-person referrals and internet advertisements, including: referral from a healthcare provider, travel clinic or immigrant/ refugee health screening, Seattle Flu Study community kiosk, school or place of work; printed flyers posted at community locations; and targeted online advertisements (e.g. Facebook, Instagram, Google).

Recruitment materials direct eligible participants to the study website (www.seattleflu.org, henceforth referenced as the “*Study Website*”). Individuals screen themselves for eligibility on the study website by providing their age, home zip code, information about presence and duration of symptoms, and confirming home internet access. Individuals are informed about the study purpose, risks, and benefits, and sign an online form to provide consent to participate in the research. Numerous consent forms are available to accommodate the age range of participants (Figure 1).

### Expected Sample Size

The study aims to enroll 3,000 participants over the course of the 2019-2020 season. We estimate 2,400 (80%) participants will return their nasal swab kit to the laboratory for respiratory pathogen testing and that 240 (10%) of specimens will be positive for influenza by molecular testing (Table 2).

**Table 2:**
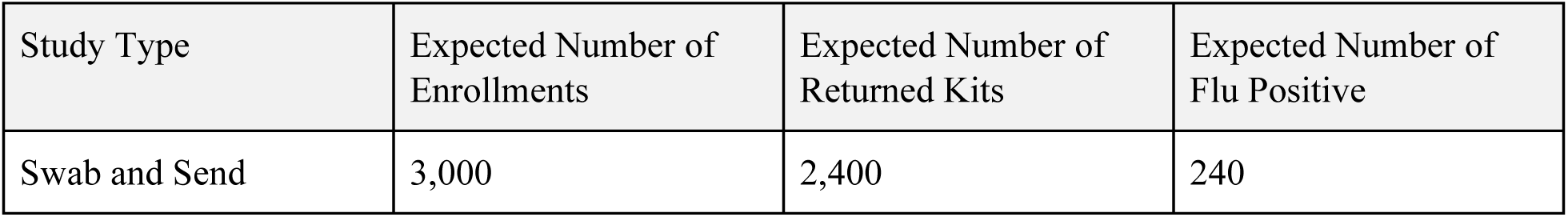
Sample size aims and expected number of influenza-positive samples

**Table 3:**
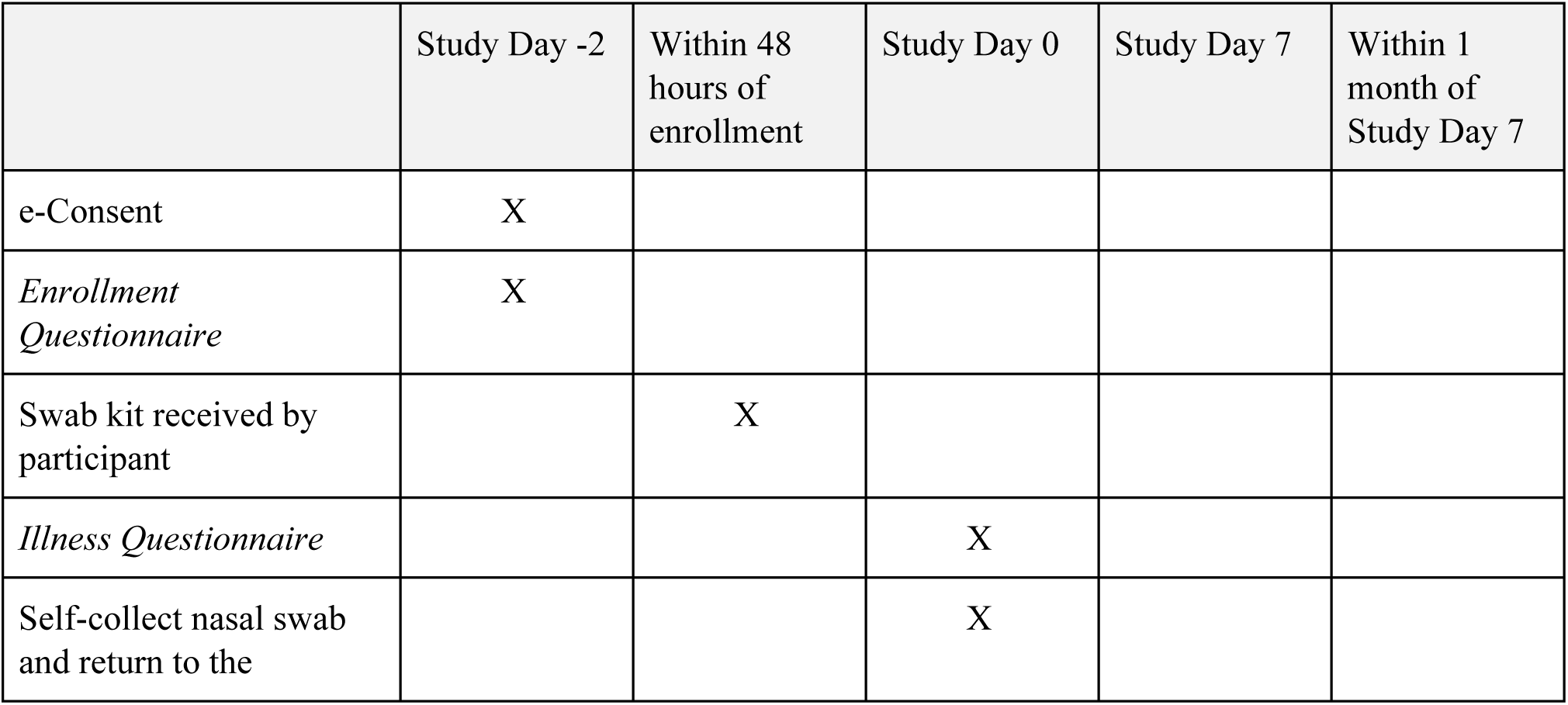
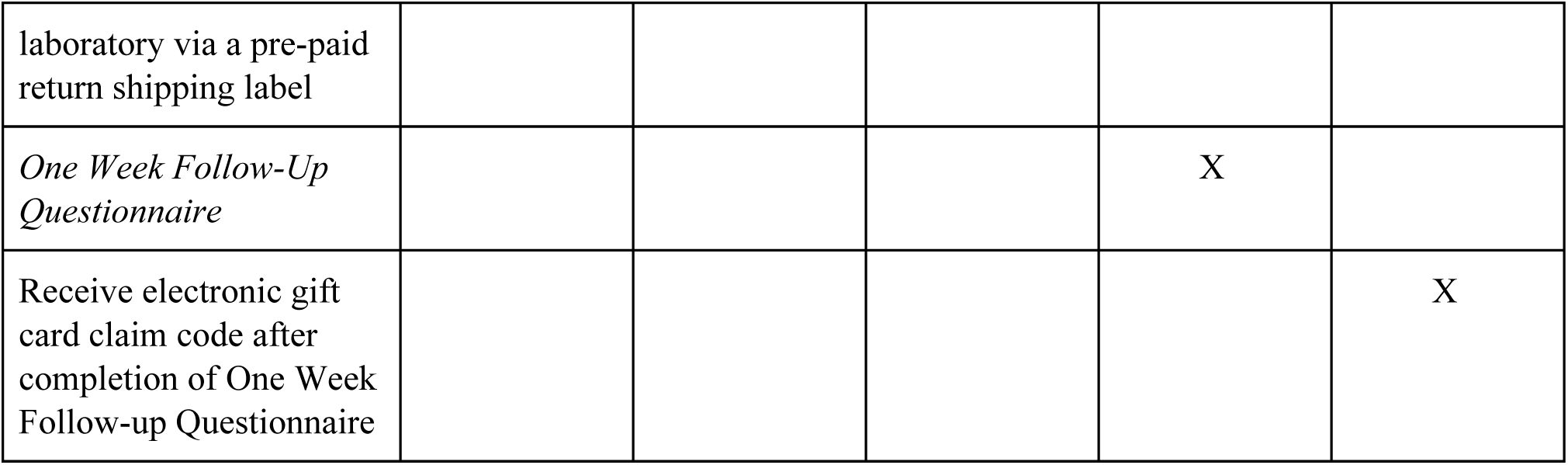
Schedule of events for each participant.

**Table 4:** REDCap Survey Instruments

| Instrument Name | Trigger for instrument | Who Completes |
| --- | --- | --- |
| Eligibility Screening (Seattle Flu Study website enrollment wizard) | Seattleflu.org connects with REDCap | Participant |
| e-Consent for Swab and Send | Seattleflu.org sorts individuals into “Swab and Send” study arm | Participant/Legally authorized representative |
| *Enrollment Questionnaire* | e-Consent completed | Participant |
| *Illness Questionnaire & Nasal Swab Collection* | Study staff ships a swab kit to the participant’s house | Participant |
| *Post Collection Data Entry and Quality Control* | Upon arrival at the University of Washington BBI Laboratory | Lab Staff |
| *One Week Follow-up Questionnaire* | Automatically generated email containing a survey link will be sent to the participant seven days after completion of the *Illness Questionnaire* | Participant |

### Outcomes

#### Primary Outcomes

1. Proportion of individuals who complete study procedures, including return of test kit and completion of questionnaire among those who test positive for respiratory pathogens
2. Usability and feasibility of self-collected nasal swabs for diagnosis of respiratory pathogens in symptomatic individuals

#### Secondary Outcomes

1. Clinical and sociodemographic characteristics of participants with respiratory pathogens who enrolled online through a home-based surveillance system and self-collected nasal swabs
2. Variation in symptom presence, duration and severity between individuals testing positive for different respiratory viruses
3. Behavioral and healthcare-seeking outcomes among individuals with and without respiratory pathogens detected, including healthcare utilization and work and school absenteeism

### Data Collection

After consent, participants complete an online *Enrollment Questionnaire*, provide their home address and contact information such as an email address or phone number. Participants are mailed a swab kit within 48 hours of submitting the Enrollment Questionnaire, which includes a *Quick Start Instruction Card*, a UTM tube (Becton, Dickinson and Company, Sparks, MD), a nylon flocked swab (COPAN Diagnostics Inc., Murietta, CA), and a return box with affixed Category B UN3373 label, as required by International Air Transport Association (IATA) guidelines[15], and a pre-paid return shipping label. Pediatric nasal swabs are available for participants 5 years of age or younger.

Upon kit receipt, participants complete an online *Illness Questionnaire* to ascertain demographics, illness characteristics, and health behaviors. At the end of the *Illness Questionnaire*, participants are prompted to self-collect a mid-nasal swab using the instructions provided on an instruction card included in the swab kit box.

Participants are instructed to place their self-collected nasal swabs directly into the UTM tube which is pre-labeled with a unique barcode. Next, participants are instructed to place the UTM tube containing the self-collected nasal swab into a specimen bag, pre-packaged with an absorbent sheet, and then place the specimen bag into the provided return shipping box. United States Postal Service (USPS) return postage and Category B UN3373 stickers are affixed to the outside of the return box. Although previous testing has demonstrated that respiratory viral RNA is stable in room-temperature UTM for up to one week[16], participants are encouraged to mail their nasal swab as soon as possible. Returned samples are estimated to arrive at the laboratory approximately 24 hours after deposit at a US Postal Service facility.

When kits are received in the study laboratory, contents of the box and errors in swab collection are recorded. Swabs are removed and viral transport media aliquoted, subjected to RNA amplification using a MagNA Pure 96 System, and tested for respiratory pathogens using TaqMan Open Array RT-PCR (Table 5)[17]. These methods are described in the Seattle Flu Study protocol[14].

**Table 5.**
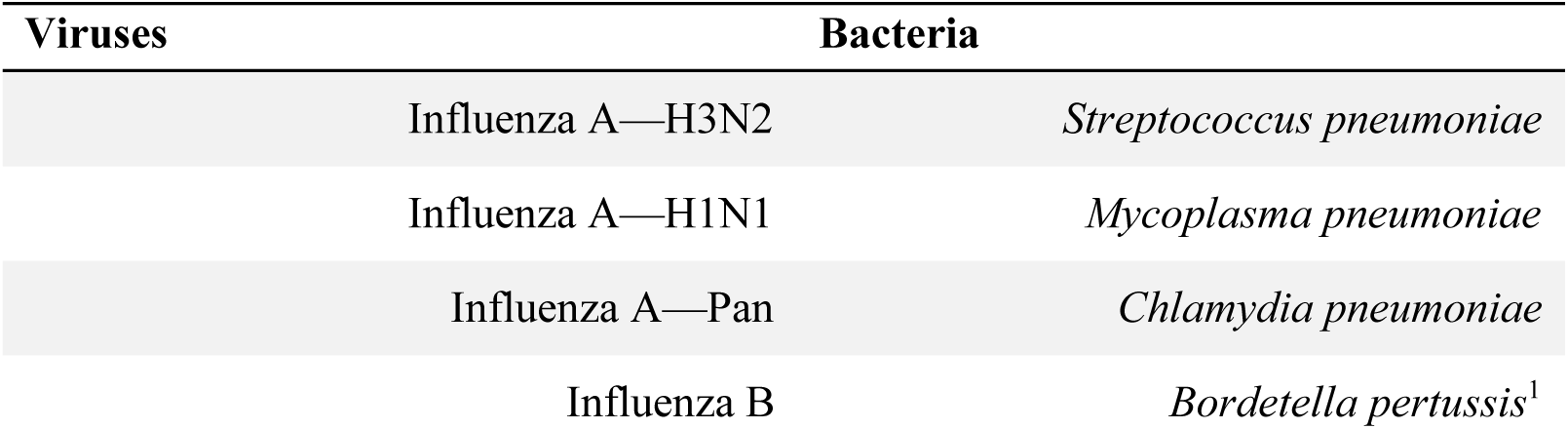
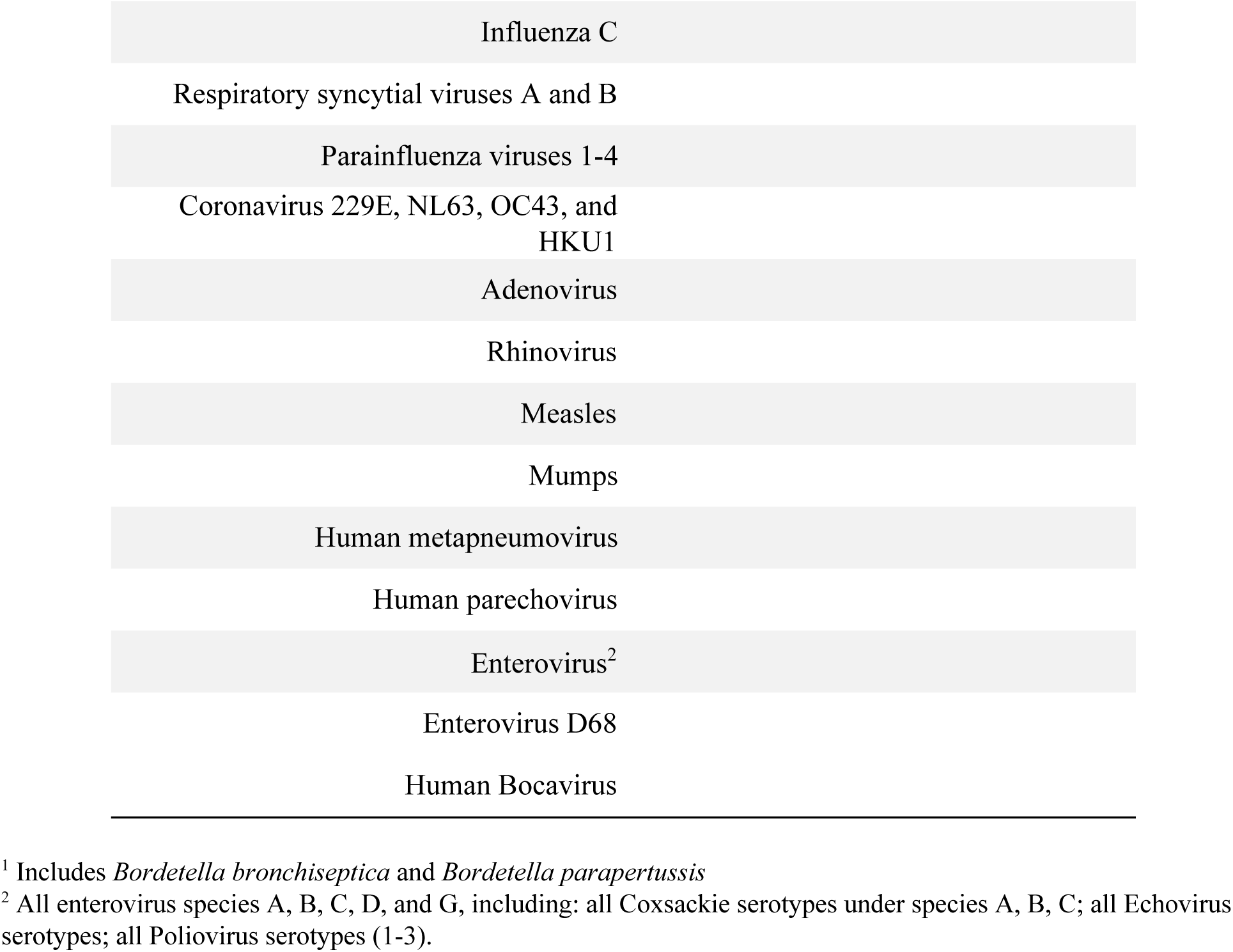
Pathogens for which all Seattle Flu Study respiratory specimens are tested using a TaqMan RT-PCR.

| Viruses | Bacteria |
| --- | --- |
| Influenza A—H3N2 | *Streptococcus pneumoniae* |
| Influenza A—H1N1 | *Mycoplasma pneumoniae* |
| Influenza A—Pan | *Chlamydia pneumoniae* |
| Influenza B | *Bordetella pertussis*<sup>1</sup> |

|  |
| --- |
| Influenza C |
| Respiratory syncytial viruses A and B |
| Parainfluenza viruses 1-4 |
| Coronavirus 229E, NL63, OC43, and HKU1 |
| Adenovirus |
| Rhinovirus |
| Measles |
| Mumps |
| Human metapneumovirus |
| Human parechovirus |
| Enterovirus<sup>2</sup> |
| Enterovirus D68 |
| Human Bocavirus |

<sup>1</sup> Includes *Bordetella bronchiseptica* and *Bordetella parapertussis*
<sup>2</sup> All enterovirus species A, B, C, D, and G, including: all Coxsackie serotypes under species A, B, C; all Echovirus serotypes; all Poliovirus serotypes (1-3).

Seven days after the nasal swab collection, participants are re-contacted to complete a *One Week Follow-Up Questionnaire* to assess the impact of their illness on behavioral outcomes such as absenteeism and healthcare-seeking behaviors (provider visits, antiviral use, etc).

### Benefits and compensation

Participants receive a $5 digital gift card for completing the *Illness Questionnaire* and returning their self-collected swab; an additional $5 gift card will be provided to participants who completed the *One Week Follow-Up Questionnaire*. Participants are able to view their test results for influenza A and B and RSV on the *Study Website* within 3 months of when the swab is collected by entering their unique sample barcode.

### Data security and privacy

Participant information is kept confidential, and questionnaire and sample data are linked to participants using a unique identification number assigned at enrollment; only the direct study team will have access to the linkage. All study questionnaires are collected through REDCap, which is Title 21 CFR Part 11 compliant[18]. Access to de-identified, aggregate study data and analysis code will be publicly available on the *Study Website*.

### Statistical Methods

Descriptive statistics of frequency/percentage and mean/standard deviation will be performed for categorical and continuous covariates, respectively. Bivariate analyses will be conducted using parametric and nonparametric tests as appropriate, with statistical significance defined as p<0.05. Respiratory pathogen prevalence is defined as the total number of cases of a pathogen detected out of the total number of tested samples. Odds ratios and 95% confidence intervals will be estimated using negative binomial or Poisson regressions with robust standard errors.

Additional analyses will occur, as described elsewhere in the Seattle Flu Study protocol[14].

## Data Availability

The data will be accessed only by authorized individuals on the study team. Access to de-identified, aggregated data and analysis code will be publicly available on the study web page (www.seattleflu.org).

## Ethics and Dissemination

Ethical approval was granted by the University of Washington IRB. Additionally, the Seattle Flu Study has a scientific advisory board to whom it reports bi-annually. These reports include updates on enrollment, preliminary results, and protocol modifications.

Our study group will present the results of the Swab and Send sub-study at national and international research conferences, through peer-reviewed publications, and on the study website. Any changes to the study protocol will be communicated to journals in accordance with that journal’s reporting policies. We will follow STrengthening the Reporting of OBservational studies in Epidemiology (STROBE) reporting guidelines, as applicable[19]. Study materials, including the *Quick Start Instruction Card*, kit design, and study questionnaires, will be available upon request with approval of the study team.

## Seattle Flu Study Investigators

### Principal Investigators

Helen Y. Chu, MD, MPH^1,2^; Michael Boeckh, MD, PhD^1,2,5^; Janet A. Englund, MD^1,2,6^; Michael Famulare, PhD^7^; Barry R. Lutz, PhD^2,8^; Deborah A. Nickerson, PhD^2,3^; Mark J. Rieder, PhD^2^; Lea M. Starita, PhD^2,3^; Matthew Thompson, MD, MPH, DPhil^9^; Jay Shendure, MD, PhD^2,3,10^; and Trevor Bedford, PhD^2,3,5^

### Co-Investigators

Amanda Adler, MS^6^; Elisabeth Brandstetter, MPH^1^; Jeris Bosua, BA^11^; Shari Cho, MS^2^; Kairsten A. Fay, BSc^5^; Chris D. Frazar, MS^2^; Chelsey Graham, MEng^2^; Peter D. Han, MS^2^; James Hadfield, PhD^2^; Misja Ilcisin, BSc^5^; Jessica Heimonen, MPH^1^; Shichu Huang, PhD^8^; Michael L. Jackson, PhD, MPH^12^; Anahita Kiavand, MS^1^; Ashley E. Kim, BS^1^; Louise E. Kimball, PhD^5^; Kirsten Lacombe, RN, MSN^6^; Jover Lee, BSc^5^; Jennifer Logue, BS^1^; Victoria Lyon, MPH^9^; Denise J. McCulloch, MD MPH^1^; Jennifer Mooney^4^; Kira L. Newman, MD, PhD^1^; Audrey Osterbind, BA^1^; Matthew Richardson, BA^3^; Thomas R. Sibley, BA^5^; Margaret M. Van de Loo, BA^4^; Monica L. Zigman Suchsland, MPH^9^; and Caitlin Wolf, BS^1^

### Affiliations

1 Department of Medicine, University of Washington

2 Brotman Baty Institute For Precision Medicine

3 Department of Genome Sciences, University of Washington

4 Formative

5 Vaccine and Infectious Disease Division, Fred Hutchinson Cancer Research Center

6 Seattle Children’s Research Institute

7 Institute for Disease Modeling

8 Department of Bioengineering, University of Washington

9 Department of Family Medicine, University of Washington

10 Howard Hughes Medical Institute

11 Blaze Clinical

12 Kaiser Permanente Washington Health Research Institute

